# Prevalence and Associated Factors of Human Tungiasis Among Primary School Children in Kamushoko Parish, Bubaare Sub-County, Mbarara District, Uganda

**DOI:** 10.64898/2026.07.03.26357206

**Authors:** Desire Conrad Inziku, Agaba Patrick, Mukoda Gloria, Ahumuza Arnold, Kitimbo Boneface Mark, Kalyetsi Rogers, Kalembe Salome

## Abstract

**Background:** Human Tungiasis is a neglected tropical skin disease caused by the sand flea Tunga penetrans, associated with pain, inflammation, secondary bacterial infections, and significant morbidity. Despite its health impact on school children, it remained understudied in Kamushoko Parish, Bubaare Sub-County, Mbarara District, Uganda.

**Objectives:** To determine the prevalence and identify associated factors of human Tungiasis among primary school children in Kamushoko Parish, Bubaare Sub-County, Mbarara District.

**Methods:** A school-based cross-sectional descriptive study was conducted among 415 primary school children (P1–P7) at Katooma II and Komuyaga Primary Schools in March 2026. A census sampling approach enrolled all eligible pupils. Data were collected through structured face-to-face questionnaires and clinical examination of exposed areas. Tungiasis diagnosis was based on clinical identification of characteristic lesions with microscopic confirmation of extracted specimens. Bivariate and multivariate logistic regression analyses identified independently associated factors.

**Results:** Of 415 children examined, 22 had active Tungiasis lesions, yielding an overall prevalence of 5.3% (95% CI: 3.53%–7.90%). Katooma II Primary School recorded a slightly higher prevalence (6.29%) than Komuyaga Primary School (4.78%), though the difference was not statistically significant (p = 0.499). Male children had higher odds of infection than females (7.1% vs. 3.7%). Children aged 9–10 years had the highest prevalence (7.7%). In the multivariate logistic regression, walking barefoot at school was the only independently significant predictor of infection (aOR = 0.18 for home vs. school; 95% CI: 0.04–0.78, p = 0.022).

**Conclusion:** The prevalence of Tungiasis among primary school children in Kamushoko Parish was 5.3%, confirming ongoing transmission in the community. Walking barefoot at school was the only independent predictor of infection, highlighting the school environment as a critical intervention site. School-based strategies including grounds improvement, routine classroom cleaning, enforcing no-barefoot policies, and health education are recommended.

## 1. Introduction

Human Tungiasis is a highly neglected tropical skin disease caused by the sand flea Tunga penetrans, the gravid female of which penetrates and embeds into the skin particularly the feet causing pain, inflammation, and significant morbidity[1]. The burden of this neglected tropical disease (NTD) is disproportionately borne by marginalized populations, particularly those living in rural areas with limited access to healthcare services and poor socioeconomic conditions [2].

In sub-Saharan Africa, systematic reviews and meta-analyses have revealed a pooled prevalence of 33.4% (95% CI: 27.6–39.8%), with some countries recording prevalence rates as high as 46.5% in Ethiopia, 44.9% in Cameroon, and 42.0% in Tanzania [3]. Uganda, like many other sub-Saharan African countries, faces a significant burden of human Tungiasis, with previous studies documenting varying prevalence rates: 22.5% in Mayuge District and up to 84.6% in some parishes in northeastern Uganda [4, 5].

Mbarara District in southwestern Uganda represents a typical rural setting where Tungiasis is likely endemic, given prevailing socioeconomic and environmental factors. Kamushoko Parish, Bubaare Sub-County, is one such area where reports of infestation exist but no systematic epidemiological study had been conducted. Preliminary observations from selected schools revealed muddy and dusty classroom floors, children walking barefoot, and poor hygiene practices as known risk factors for T. penetrans transmission [6].

This study therefore sought to determine the prevalence of human Tungiasis among primary school children in Kamushoko Parish and to identify specific environmental, behavioral, and socioeconomic factors associated with disease occurrence. The findings provide essential evidence for designing targeted interventions and informing school health policies in rural Mbarara District.

## 2. Materials and Methods

### 2.1 Study Design and Setting

A school-based cross-sectional descriptive study was conducted in March 2026 at two primary schools, Katooma II Primary School (n = 143 pupils) and Komuyaga Primary School (n = 272 pupils) in Kamushoko Parish, Bubaare Sub-County, Mbarara District, southwestern Uganda. The study site was selected based on preliminary reports of Tungiasis cases and the presence of environmental risk conditions [6].

### 2.2 Study Population and Sampling

The study employed a census sampling approach, enrolling all eligible Primary One to Primary Seven (P1–P7) pupils in both schools (total n = 415). Nursery pupils were excluded because the structured questionnaire required basic literacy. Pupils who were absent during data collection or whose school administrators refused consent were also excluded.

### 2.3 Data Collection

Data were collected through structured face-to-face questionnaires administered in both English and Runyankore (local language). The questionnaire captured socio-demographic characteristics, knowledge of Tungiasis, behavioral practices (footwear use, hygiene), and environmental factors (floor type, compound cleanliness). The questionnaire was pre-tested outside the study area prior to implementation.

All participants underwent thorough clinical examination of the feet, hands, and other exposed areas for Tungiasis lesions. Lesions were classified according to the Fortaleza classification as low (below 5 lesions), moderate (5–10), or high (≥10) severity. Specimens extracted from confirmed cases were preserved in 10% formalin and prepared as permanent mounts for microscopic confirmation using the DPX mounting technique.

### 2.4 Case Management

Children with confirmed Tungiasis received basic care including flea extraction using sterile safety pins and application of coconut oil-based Vaseline. Severe cases were referred to Bubaare Health Centre III. Health education on Tungiasis prevention was provided to all participants and school staff.

### 2.5 Data Analysis

Data were entered and cleaned in an electronic database. Prevalence was estimated with 95% confidence intervals (CI). Chi-square tests assessed associations between categorical variables and Tungiasis status. Variables with a p-value < 0.2 in bivariate analysis were entered into a multivariate logistic regression model to identify independently associated factors. Statistical significance was set at p < 0.05.

### 2.6 Ethical Considerations

Ethical approval was obtained from the Faculty Research Committee of Mbarara University of Science and Technology. Administrative clearance was obtained from Bubaare Sub-County local authorities. For all participants aged 5–18 years, assent was obtained from the children and consent from school administration and teachers. Participation was voluntary and confidential, with unique study codes replacing personal identifiers.

## 3. Results

### 3.1 Sociodemographic Characteristics

A total of 415 primary school children participated in this study. The majority were aged 11–18 years (48.4%), while 7.5% were aged 5–6 years. Slightly more females (52.5%) than males (47.5%) participated. Most participants were in the lower classes (P1–P3, 49.6%). Two-thirds of participants were from Komuyaga Primary School (65.5%). Sociodemographic characteristics are summarized in Table 1.

**Table 1:** Sociodemographic characteristics.

| Variable | Frequency (n=415) | Percentage (%) |
| --- | --- | --- |
| **Age (years)** |  |  |
| 5-6 | 31 | 7.5% |
| 7-8 | 80 | 19.3% |
| 9-10 | 103 | 24.8% |
| 11-18 | 201 | 48.4% |
| **Gender** |  |  |
| Male | 197 | 47.5% |
| Female | 218 | 52.5% |
| **Class** |  |  |
| P1 – P3 | 206 | 49.6% |
| P4 – P5 | 144 | 34.7% |
| P6 – P7 | 65 | 15.7% |

Table 1: Sociodemographic characteristics
| **School** |  |  |
| --- | --- | --- |
| Katooma II Primary School | 143 | 34.5% |
| Komuyaga Primary School | 272 | 65.5% |

### 3.2 Prevalence of Human Tungiasis

Of the 415 children examined, 22 had active Tungiasis lesions, yielding an overall prevalence of 5.3% (95% CI: 3.53%–7.90%). The prevalence at Katooma II Primary School was 6.29% (95% CI: 3.35%– 11.53%) and at Komuyaga Primary School was 4.78% (95% CI: 2.81%–8.00%). The most affected sites were the toes and fingers.

### 3.3 Distribution of Tungiasis by Sociodemographic Characteristics

Male children had a higher Tungiasis prevalence (7.1%) compared to females (3.7%). Children aged 9–10 years had the highest prevalence (7.7%), while those aged 5–6 years had the lowest (3.2%). Among class levels, P1–P3 pupils recorded the highest prevalence (6.25%). None of these differences reached statistical significance. The distribution of Tungiasis by sociodemographic characteristics is presented in Table 2.

**Table 2:** Distribution of Tungiasis by Sociodemographic characteristics.

| Variables | Total (n%) | Tungiasis status |  | p-value |
| --- | --- | --- | --- | --- |
|  |  | Positive (%) | Negative (%) |  |
| School |  |  |  |  |
| Katooma II Primary School | 143 (34.5%) | 9 (6.29%) | 134 (93.71%) | 0.499 |
| Komuyaga Primary School | 272 (65.5%) | 13 (4.78%) | 259 (95.22%) |  |
| Class |  |  |  |  |
| P1 – P3 | 208 (50.12%) | 13 (6.25%) | 195 (93.75%) | 0.677 |
| P4 – P5 | 144 (34.70%) | 6 (4.17%) | 138 (95.83%) |  |
| P6 – P7 | 63 (15.18%) | 3 (4.76%) | 60 (95.24%) |  |
| Sex |  |  |  |  |
| Male | 197 (47.5%) | 14 (7.1%) | 183 (91.8%) | 0.119 |
| Female | 218 (52.5%) | 8 (3.7%) | 210 (96.3%) |  |

| **Age group** |  |  |  |  |
| --- | --- | --- | --- | --- |
| 5 – 6 years | 31 (7.5%) | 1 (3.2%) | 30 (96.8%) | 0.580 |
| 7 – 8 years | 81 (19.3%) | 3 (3.7%) | 78 (96.3%) |  |
| 9 – 10 years | 103 (24.8%) | 8 (7.7%) | 95 (92.3%) |  |
| 11 – 18 years | 200 (48.4%) | 10 (5%) | 190 (95%) |  |

### 3.4 Bivariate and Multivariate Analysis of Associated Factors

Male children had twice the odds of having Tungiasis compared to their female counterparts and children aged 9–10 years had the highest odds of Tungiasis compared to the reference group of 11–18 years. Regarding class level, children in P1 – P3 had slightly higher odds of Tungiasis compared to the reference group of Primary Six to Primary Seven. The place where children most commonly walk barefoot emerged as the most important and statistically significant factor in this study. Using school as the reference category, children who primarily walk barefoot at home had significantly lower odds of Tungiasis in both the bivariate and multivariate analyses. This association remained robust and statistically significant. Results are presented in Table 3.

**Table 3:** Bivariate and Multivariate Analysis of Associated Factors.

| **Variables** | **Bivariate** |  | **Multivariate** |  |
| --- | --- | --- | --- | --- |
|  | **cOR(95%CI)** | **P-value** | **aOR(95%CI)** | **P-value** |
| **Gender** |  |  |  |  |
| Male | **2.01 (0.82 – 4.90)** | **0.125** | 1.57 (0.62 – 4.01) | 0.342 |
| Female | 1.00 |  | 1.00 |  |
| **Age group** |  |  |  |  |
| 5 – 6 years | 0.63 (0.08 – 5.13) | 0.669 | — | — |
| 7 – 8 years | 0.73 (0.20 – 2.73) | 0.641 | — | — |
| 9 – 10 years | 1.60 (0.61 – 4.19) | 0.338 | — | — |
| 11 – 18 years | 1.00 |  |  |  |
| **Class** |  |  |  |  |
| P.1 – P.3 | 1.33 (0.37 – 4.84) | 0.662 | — | — |
| P.4 – P.5 | 0.87 (0.21 – 3.59) | 0.847 | — | — |
| P.6 – P.7 | 1.00 |  |  |  |
| **Floor type at home** |  |  |  |  |
| Earthen | 0.60 (0.21 – 1.65) | 0.318 | — | — |
| Cement | 1.00 |  |  |  |
| **Footwear usage** |  |  |  |  |
| Always | 1.00 |  | 1.00 |  |
| Sometimes | **2.89 (0.66 – 12.66)** | **0.159** | 2.34 (0.52 – 10.60) | 0.271 |
| Never | 1.12 (0.10 – 12.76) | 0.926 | 1.20 (0.10 – 13.99) | 0.882 |
| **Cleaning of the compound** |  |  |  |  |
| Daily | 1.00 |  |  |  |
| Weekly | 1.25 (0.35 – 4.45) | 0.728 | — | — |
| Rarely | 0.66 (0.15 – 2.95) | 0.588 | — | — |
| **Place walked barefoot** |  |  |  |  |
| Home | **0.17 (0.04 – 0.72)** | **0.016** | **0.18 (0.04–0.78)** | **0.022** |
| Farm | 0.57 (0.15 – 2.18) | 0.409 | 0.54 (0.14–2.12) | 0.376 |
| School | 1.00 |  | 1.00 |  |

## 4 Discussion

### 4.1 Prevalence of Tungiasis

The overall Tungiasis prevalence of 5.3% (95% CI: 3.53%–7.90%) observed in this study was lower than most estimates reported across Uganda and the broader East African region. Within Uganda, the pooled prevalence stands at 20.1% [3], with school-based studies reporting 17.3% in Butaleja District [4]. Comparable school-based studies across East Africa documented 48% in Kilifi County, Kenya [7], 31.2% among primary school children in Zanzibar [8], and 28.5% in Bugesera District, Rwanda [9]. The present finding is more consistent with estimates from southwestern and western Uganda, Bushenyi District (12.8%) and Wakiso District (9.2%) suggesting a regional pattern of comparatively lower transmission in Uganda’s southwestern highland zone, attributable to cooler temperatures, relatively better housing standards, and reduced sand flea survival [2].

### 4.2 Distribution by Sex and Age

Male children recorded a higher prevalence (7.1%) compared to females (3.7%), consistent with evidence from Nigeria [10] and Kenya [7], where gender-differentiated activity patterns and less consistent footwear use among boys were cited as explanatory factors. Children aged 9–10 years had the highest prevalence (7.7%), consistent with Feldmeier’s observation that children aged 5–14 years bear the highest Tungiasis burden due to frequent barefoot walking, prolonged outdoor play, and thinner skin permitting easier flea penetration [2]. The relatively lower prevalence in the youngest age group (5–6 years) likely reflects greater parental supervision, while the slight decline in older children (11–18 years) may reflect growing awareness and more consistent footwear use.

### 4.3 The School Environment as the Primary Site of Transmission

The most important finding of this study was the statistically significant association between walking barefoot at school and Tungiasis infection (aOR = 0.18, 95% CI: 0.04–0.78, p = 0.022), which was the sole independently significant factor after multivariate adjustment. This indicates that the school environment, characterized by sandy, poorly maintained grounds and uncemented or unkept classroom floors constitutes the primary site of T. penetrans exposure for children in Kamushoko Parish. Feldmeier noted that T. penetrans thrives best in dry, sandy soils commonly found on unimproved grounds of rural primary schools in sub-Saharan Africa [2]. The non-significant finding for household floor type in this study further supports this interpretation, suggesting that home-based transmission is secondary to school-based exposure in this population.

Footwear usage showed a trend in the expected protective direction (aOR = 2.34 for sometimes vs. always wearing shoes), though this did not reach statistical significance after adjustment, likely due to the small number of confirmed cases limiting statistical power. Feldmeier cautioned that footwear use as a preventive measure faces practical barriers in resource-poor communities, including cost and cultural norms of removing shoes indoors [2].

### 4.4 Teachers’ Perspectives

Near-universal teacher knowledge of Tungiasis (88.9%) and unanimous recognition of its impact on learning reinforce the community-level awareness of the disease. However, one-third of teachers reported having no defined response protocol for infected pupils, mirroring institutional gaps identified across school-based studies in Uganda and East Africa [5, 11]. The dominant challenge identified by teachers of parental ignorance and negligence aligns with findings from Mutebi et al. that household-level knowledge deficits perpetuate transmission cycles even when school-level awareness is relatively high [12].

## 5. Conclusion

The prevalence of Tungiasis among primary school children in Kamushoko Parish was 5.3%, which was low but confirming ongoing transmission at the schools and in the community. Walking barefoot was the only independent predictor of infection, highlighting the school environment as a critical site for intervention. The infection rates were uniformly distributed across both schools, all class levels, age groups, and sexes, demonstrating that all children in the parish share the same risks of infection. The school environment was the single, independently confirmed site of Tungiasis transmission in this population.

## 6. Recommendations

Given the confirmed Tungiasis burden in Kamushoko Parish, the school authorities should enforce a strict no-barefoot policy and prioritize improvement of school grounds. Universal health education covering transmission, prevention, and symptom recognition should be delivered to all pupils and teachers across both schools, since no subgroup is exempt from risk.

## Data Availability

All data regarding this studies is readily available and will be provided upon request

## 7. Declarations

### Ethical Approval and Consent

Ethical approval was obtained from the Faculty Research Committee of Mbarara University of Science and Technology (Ref: MUST/MLS/023). Assent was obtained from child participants; consent was obtained from school administration and teachers.

### Authors’ Contributions

All five authors contributed equally to study conceptualization, data collection, analysis, and manuscript preparation.

## Acknowledgements

The authors thank supervisors Mr. Kalyetsi Rogers and Mrs. Kalembe Salome for their guidance. Gratitude is extended to the administration and pupils of Katooma II and Komuyaga Primary Schools, the leadership of Bubaare Sub-County, and the staff of Bubaare Health Centre III.

## References

1. Sánchez-Cárdenas, C.D., et al., Tungiasis, in Skin Disease in Travelers. 2024, Springer. p. 257–265.

2. Feldmeier, H., et al., Tungiasis—a neglected disease with many challenges for global public health. PLoS neglected tropical diseases, 2014. 8(10): p. e3133.

3. Obebe, O.O. and O.O. Aluko, Epidemiology of tungiasis in sub-saharan Africa: a systematic review and meta-analysis. Pathogens and global health, 2020. 114(7): p. 360–369.

4. Wafula, S.T., et al., Prevalence and risk factors associated with tungiasis in Mayuge district, Eastern Uganda. The Pan African medical journal, 2016. 24: p. 77.

5. McNeilly, H., et al., Reduction of tungiasis prevalence, intensity, and morbidity during a two-year long community-based tungiasis control project in a hyperendemic region in Karamoja, Uganda. PLOS Neglected Tropical Diseases, 2025. 19(6): p. e0013149.

6. Adriko, M., The Prevalence and Risk Factors Associated with Tungiasis Infestations in Uganda: Implications for Vector Borne and Neglected Tropical Disease Control, in Zoonosis of Public Health Interest. 2022, IntechOpen.

7. Elson, L., et al., Prevalence, intensity and risk factors of tungiasis in Kilifi County, Kenya II: Results from a school-based observational study. PLoS neglected tropical diseases, 2019. 13(5): p. e0007326.

8. Mtunguja, M., et al., Tungiasis infection among primary school children in Northeastern Tanzania: prevalence, intensity, clinical aspects and associated factors. IJID regions, 2023. 7: p. 116–123.

9. Nsanzimana, J., et al., Factors associated with tungiasis among primary school children: a cross-sectional study in a rural district in Rwanda. BMC public health, 2019. 19(1): p. 1192.

10. Ugbomoiko, U.S., et al., Prevalence and clinical aspects of tungiasis in south-west Nigerian schoolchildren. Tropical doctor, 2017. 47(1): p. 34–38.

11. McNeilly, H., et al., Tungiasis stigma and control practices in a hyperendemic region in Northeastern Uganda. Tropical Medicine and Infectious Disease, 2023. 8(4): p. 206.

12. Mutebi, F., et al., Animal and human tungiasis-related knowledge and treatment practices among animal keeping households in Bugiri District, South-Eastern Uganda. Acta tropica, 2018. 177: p. 81–88.

